# Explainable machine learning on weighted connectivity networks across frequencies for outcome prediction in comatose patients

**DOI:** 10.1101/2025.11.21.25340725

**Authors:** Arthur Verdeyme, Jake P. Grainger, Marzia De Lucia, Sofia C. Olhede

**Affiliations:** Institute of Mathematics, Ecole Polytechnique Fédérale de Lausanne, Lausanne, Switzerland; Brain-Body and Consciousness Laboratory, Department of Clinical Neuroscience, Lausanne University Hospital, University of Lausanne, Lausanne 1011, Switzerland; Center for Biomedical Imaging, Lausanne 1011, Switzerland

**Author notes:** These authors contributed equally to this work (Marzia De Lucia, Sofia C. Olhede). These authors also contributed equally to this work. Current Address: Dept/Program/Center, Institution Name, City, State, Country. Deceased. Membership list can be found in the Acknowledgments section.

## Abstract

**Background:** Accurate early prediction of neurological outcomes in comatose patients after cardiac arrest is critical for guiding therapeutic decisions and improving individualized care. Current electroencephalographic (EEG) approaches typically rely on threshold-based or binarized measures of functional connectivity, which may overlook key subtleties of brain dynamics and limit clinical interpretability. We aimed to develop an explainable machine learning framework using weighted EEG connectivity and topological network features to improve early outcome prediction after cardiac arrest.

**Methods and Findings:** We analyzed EEG recordings from comatose patients within the first 24 hours after cardiac arrest, recruited from multiple intensive care units across Switzerland. Weighted functional connectivity networks were computed using the debiased weighted phase-lag index (dwPLI) across 1–40 Hz. We extracted multi-frequency topological metrics describing global integration and local segregation, and used these as features in an explainable machine learning classifier to predict long-term neurological outcome. Model interpretation was performed using Shapley additive explanations (SHAP). The classifier achieved up to 95% accuracy in distinguishing patients with favourable versus unfavourable outcomes, matching or surpassing existing approaches. SHAP analyses identified delta-band features, particularly path length and clustering coefficient, as the most informative predictors, highlighting differences in large-scale integration and local segregation between outcome groups. Main limitations include the moderate cohort size and the dependency on expert-guided artefact rejection during EEG preprocessing.

**Conclusions:** Our findings demonstrate that weighted network topology and multifrequency EEG analysis provide valuable, interpretable biomarkers of coma outcome. The proposed explainable AI framework offers a transparent, quantitative, and clinically meaningful approach for early neurological prognostication after cardiac arrest and may inform the design of future decision-support tools in critical care neurology.

**Author summary:** After a cardiac arrest, many patients remain unconscious in intensive care. Families and clinicians face difficult decisions, yet early and reliable signs of recovery are scarce. In this study, we analyze routine bedside EEG recordings by viewing the brain as a network. We do not study individual connections. Instead, we transform the data into network topological features, simple summary measures of the network’s overall shape and organization. Using these features across a range of slower and faster brain rhythms, we train a transparent prediction model to estimate each patient’s chance of regaining consciousness. For each patient, the model indicates which network features and which rhythm ranges most influenced its estimate, in clear figures that clinicians can examine. We find that these network summaries add useful information to current bedside assessments and may help structure earlier, more consistent conversations about prognosis. We present a step-by-step analysis designed with clinical workflows in mind. Our goal is to support, not replace, medical judgment, and to help build explainable tools for disorders of consciousness.

## Introduction

Predicting the neurological outcomes of comatose patients after cardiac arrest is a critical task as they influence treatment decisions and the allocation of clinical resources. Electroencephalography (EEG) has been widely used in this setting because it is portable, cost-effective, and suitable for bedside monitoring [1, 2, 3, 4]. In particular, networks that represent the functional connectivity derived from such EEG data have highlighted how the brain is modularly organised, with interconnected hubs that transfer information when consciousness is in a normal state [5, 6, 7]. However, in patients with disorders of consciousness, these connectivity patterns generally break down, affecting the effectiveness of information transfer [8]. In such patients, the features obtained from functional EEG connectivity have been studied to assess clinical severity and predict long-term outcomes [9, 10].

Most existing methods rely on binarised functional connectivity networks [9], black-box machine learning (ML) models [11, 12], or band-specific analysis [13], which limit both accuracy and interpretability in real-world settings. Yet these systems face obstacles in clinical adoption—lack of robustness, task complexity, and the high stakes of patient care can undermine clinician confidence unless the model has good predictive performance and its behaviour is made transparent [14]. In fact, understanding why a model makes a certain prediction is crucial to developing trust [15, 16, 17, 18] and can even improve the prediction itself [19, 20, 21, 22, 23]. Thus, bridging the gap between algorithmic output and clinical reasoning is beneficial and requires model explanations [14]. Additionally, binarising connectivity may improve signal-to-noise ratios [24] but can also mask clinically relevant information or lead to inconsistent prediction performance [25, 26, 27].

To address these aforementioned limitations, we propose a unified explainable ML framework that integrates explainable AI techniques into a statistical pipeline, utilises weighted EEG connectivity to preserve continuous connection strengths [28], and integrates multi-frequency analysis (1–40 Hz) to extract richer topological features. First, we embed Shapley-based explainability [29, 30, 31] directly into our statistical pipeline, mitigating the black-box effect and its inherent limited information on how predictions are made in ML [32]. This approach not only improves robustness but also provides patient-specific explanations of the decision-making process, effectively offering a “statistical diagnosis” for each patient.

Then, we extract topological features from weighted functional connectivity networks in contrast to binarised methods. Doing so, we prevent common issues related to such methods like the selection of optimal thresholds which may inadvertently remove important topological features from predictions [33, 34, 35]. Therefore, although binarised networks may simplify brain interactions during coma and miss some nuances, our approach keeps more of the continuous connectivity information, which is rarely done [12, 36].

To further improve predictions, our topological features are derived from different frequencies, ranging from 1 to 40Hz, using the debiased weighted phase-lag index of [37]. Hence, we include different brain functions, ultimately capturing a wider spectrum of functional connectivity to enhance performance and interpretability [38, 39, 40, 41]. In fact, multiple-frequency analyses can present a more complete understanding of the functional state of the brain, as shown in [38], [39], and [40]. By computing a set of topological features from these weighted networks [42, 43, 44, 45] and integrating information across multiple frequencies [11, 12], our aim is to improve the accuracy of predicting patient outcomes.

Our statistical pipeline includes a systematic feature selection step that removes correlated features to further reduce complexity and prevent redundancy of the model. Consequently, this also improves the reliability of our explainable framework. In fact, highly correlated features receive approximately the same importance value in the Shapley-based method [46]. However, the high correlation implies that one of the two features is redundant for prediction. Thus, removing correlated features gives a more accurate representation of the impact that a feature has on predictions and, for patients, on the decision process. We further optimise our pipeline by performing grid search hyperparameter selection to extract the ”best predictive” frequencies and bands if selected by the user of our method, the best hyperparameters of the classifier model we use and the best correlation threshold for the feature selection step.

In this paper, our aim is to address these challenges, namely, to implement an explainable pipeline for the weighted network and their different frequency information. Our combined approach represents a significant advancement over existing models by combining weighted network topology, multi-frequency analysis, and explainable AI. Not only does it have high accuracy, but it also provides simpler models while offering a transparent and interpretable alternative that is needed for adoption in critical care settings.

## Materials and methods

### Patient description and clinical assessments

This study uses an existing data set of electroencephalographic (EEG) recordings collected from 138 adult comatose patients in the first 24 hours after cardiac arrest, as previously described in detail by [13]. Each patient routinely received targeted temperature management at 33 ° C or 36 ° C. Sedation (including midazolam, propofol, and fentanyl) and neuromuscular blocking agents were administered as needed to control pain and shivering. Sixty-three channel resting-state EEGs were recorded while patients were clinically evaluated using standard neurological exams (such as the Glasgow coma scale [47] and the FOUR score [48]), and extensive follow-up evaluations determined the outcome of each patient at 3 months of postcardiac arrest. According to current guidelines, the classification of results distinguished between favourable outcome (CPC 1-2), labelled FO, and unfavourable outcome (CPC 3-5) and no patients who remain vegetative at 3 months. Thus, this data set provides a rich source of electrophysiological signals captured during a critical time window in early coma, enabling the investigation of large-scale functional connectivity patterns and their prognostic value. Additionally, in the EEG recordings from [13], some were excluded after preprocessing due to diffuse artefacts (either muscle or machine noise); reducing the number of patients to 93. In this study, patients with FO are associated with the positive class, while patients with UO are associated with the negative class.

### Ethics statement

Patients were recruited from multiple intensive care units across Switzerland, such as the University Hospitals of Lausanne (n = 69), Sion (n = 4), Fribourg (n = 2) and Bern (n = 63) between July 2014 and January 2018. The study was approved by the ethics committees of the respective institutions (Commission cantonale d’éthique de la recherche sur l’être humain du Canton de Vaud, CER-VD, approval PB 2016-00530, 23/05). Informed consent was obtained for all patients in accordance with the ethical approval.

### Electroencephalography data analysis

As a connectivity metric, we use the debiased weighted Phase-Lag Index (dwPLI) to assess phase synchronisation in the presence of volume conduction, acting as a noise and artefact resistant index [37]. Estimation of undirected connectivity was performed using Fieldtrip [49] and the dwPLI was computed across a wide frequency range from 1 to 40 Hz, in 1 Hz steps, using a Slepian tapering sequence for spectral smoothing.

Unlike conventional methods that threshold and binarise connectivity matrices [50, 9, 13, 51, 52, 53], we kept the matrices as weighted networks [42]. This approach preserves the original strength of the connections, providing a more detailed representation of neural interactions. Although binarising weighted networks can improve the signal-to-noise ratio [24], this process may lead to a loss of information and prediction performance [25]. Attempts to find optimal thresholds often involve fixing the value of certain topological features such as degrees [33, 54, 55, 56, 57], edge density [34], or clustering coefficient [35, 58], essentially removing those features from predictions. Another common approach is to select a range of thresholds to test. However, this method has significant drawbacks: the number of candidate thresholds is arbitrary, the networks are not independent samples [59], and the network summary statistics can be unstable between thresholds [60], leading to contradictory results in the literature [26].

Using weighted networks, we take advantage of the continuous nature of connectivity data to gain deeper insights into functional brain networks [61]. As such, we avoid the loss of information inherent in thresholding and binarising, allowing for a more nuanced analysis of network architecture and dynamics over time. This approach aligns with previous studies that have used weighted networks to capture the complexity of brain interactions [12, 36, 62]. The dwPLI is an estimator for an underlying positive quantity [37], however, the debiasing step can produce slightly negative weights. Since many network summary statistics require non-negative weights, we set these negative weights to zero before performing the analysis. Using weighted networks across different frequencies allows us to capture a wide spectrum of functional connectivity, enhancing both performance and interpretability.

### Network topological features

To simplify the high-dimensionality of EEG connectivity data, we use network statistics referred to as topological features, computed for every frequency from 1 Hz to 40 Hz. The key features evaluated include the clustering coefficient, characteristic path length, modularity, and participation coefficient [50, 9, 13]. In our network model, nodes represent electrodes and weighted edges represent the dwPLI values.

Going from traditional binary representations of EEG connectivity matrices to weighted graphs demands adapted definitions (Supporting Information, S1 Text) [63, 43, 44, 45]. Feature formulas relied either on node degree or inter-node distance. In weighted graphs, a node’s degree equals the sum of incident weights, reflecting interaction strength. Because dwPLI is a similarity measure, higher weights imply stronger synchrony. Therefore, distances were taken as the inverse of weights, so stronger synchrony yielded shorter functional paths [44, 64]. This conversion into a distance matrix is consistent with the actual representation of the information transfer and was adapted for features such as path length, efficiency, and communicability.

In addition to the commonly used features mentioned previously, we incorporated additional topological features that are particularly relevant in the context of weighted networks: global efficiency, edge density, and communicability. The Supporting Information (S1 Table) lists definitions and references of the seven topological features considered in this study. Together, these metrics provide a multifaceted characterisation of weighted networks, giving a more detailed description of brain functional connectivity. Considering weighted networks instead of their thresholded-binarised counterparts reduces loss of information; thus improving our identification of key network alterations associated with different cognitive states or neurological conditions.

### Statistical methodology

A statistical pipeline (Figure 1) quantified the predictive value of each topological feature and their combinations over 1–40 Hz. To reduce dimensionality while reflecting neurological functions, features were also grouped into delta (1–4 Hz), theta (5–7 Hz), alpha (8–12 Hz), beta (13–30 Hz), and gamma (*>*30 Hz) bands.

**Fig 1.**
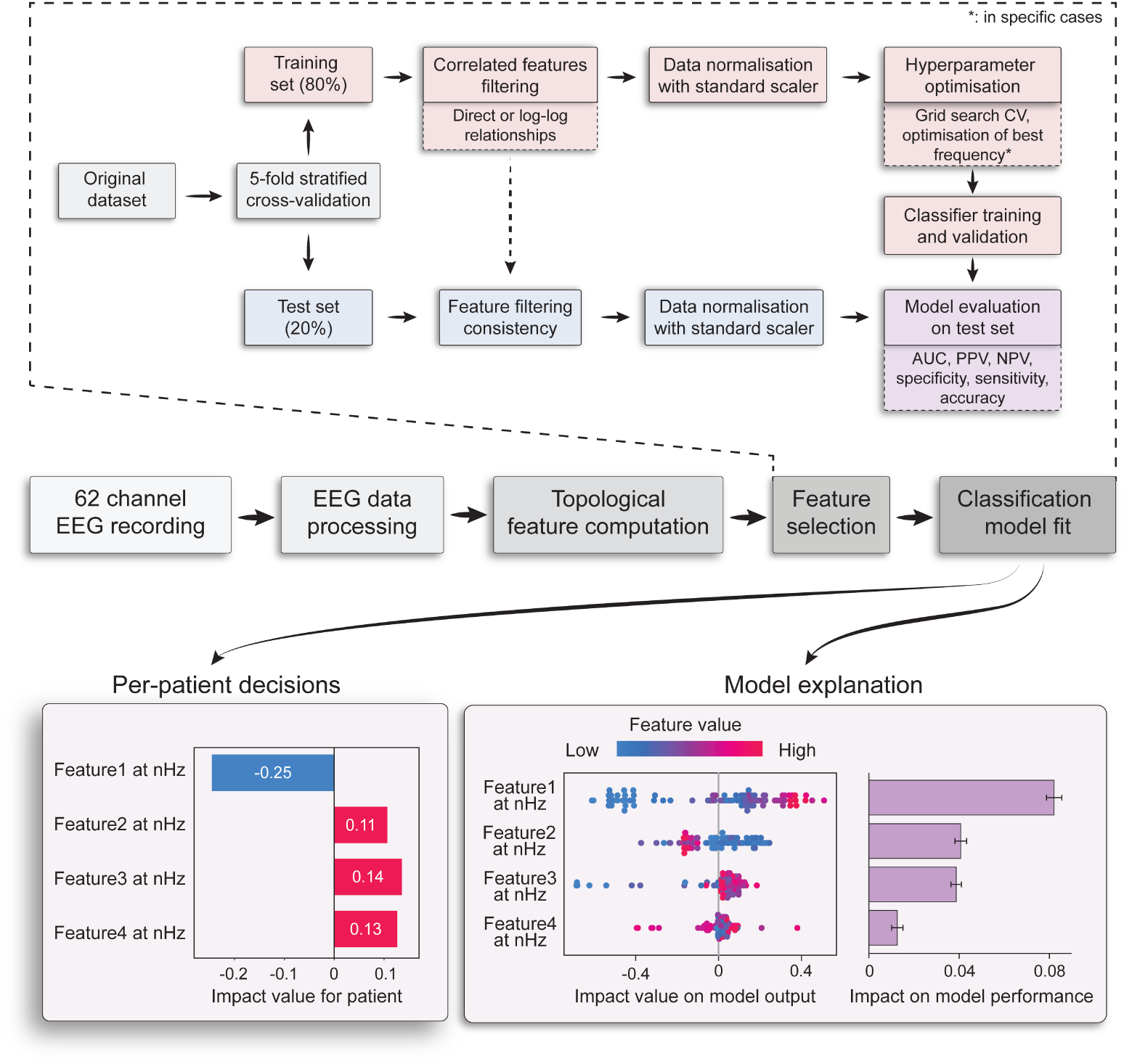
Diagram showing the complete process the data undergo when combining features for prediction, as well as the procedure to create the classifiers. This includes the acquisition of the data, its cleaning and preprocessing from [13], the computations of the weighted topological features, the correlated feature filtering, the training of the classifiers, selection of the best hyperparameters, the performance evaluation of the classifiers at the test set, and an illustrative example of an explanation of the model.

Because the cohort was imbalanced (52 FO, 41 UO), patients were stratified before an 80/20 train–test split (5-fold, four for the train set, one set aside for the test set), preserving class ratios. In the training set, features showing pairwise correlation in linear or log-log space were discarded given a correlation threshold treated as a hyperparameter to be optimised in the pipeline. This step accounts for potential linear or power-law relationships that might exist among network topological features, as described in [65], [66] and [67], cut redundancy and improve interpretability.

After feature selection, we scale the features to standardise their ranges and then fit a classifier to predict patient outcomes based on the selected features. Hyperparameters were tuned with a grid search and 4-fold stratified cross-validation that maximised the geometric mean of accuracy and sensitivity, balancing overall performance with minority-class detection. The best model was retrained on all training data and evaluated on the test set with accuracy, sensitivity, specificity, Area Under the Receiver Operating Characteristic Curve (AUC), Positive Predictive Value (PPV) and Negative Predictive Value (NPV).

To assess the statistical significance of the model performance, we performed a permutation test on the patient’s labels and extracted its associated p-value. This involves comparing the model’s performance (here the metrics mentioned just before) against a distribution of performances obtained by randomly shuffling the outcome labels and retraining our model (20000 times in our case), thereby validating that the observed results are not due to chance. As accuracy governed model selection and correlates highly with the other metrics, we chose to apply the permutation test only to accuracy. When comparing metrics across different models or single features, we further control the obtained p-values by applying a false discovery rate (FDR) correction [68]. We first explore the predictive capabilities of individual topological features and then the combinations thereof as described in the next subsection.

#### Feature-set construction

We first explored each topological feature as a stand-alone predictor within every frequency band, using a 50/50 stratified train–test split. Treating one variable at a time clarifies its isolated predictive power and reveals frequency-specific strengths (Fig. 2). As such, these exploratory results indicated that combining features could capture inter-frequency synergies. We therefore constructed the following datasets:

*TF_f_* (*f* = 1, …, 40): seven features at a single frequency *f*.
*TF*_1:40_: all seven features from every frequency (7 × 40 = 280).
*TF_δ_*, *TF_θ_*, *TF_α_*, *TF_β_*, *TF_γ_*: band-averaged features.
*TF_δ:γ_*: concatenation of the five band sets (7 × 5 = 35).

**Fig 2.**
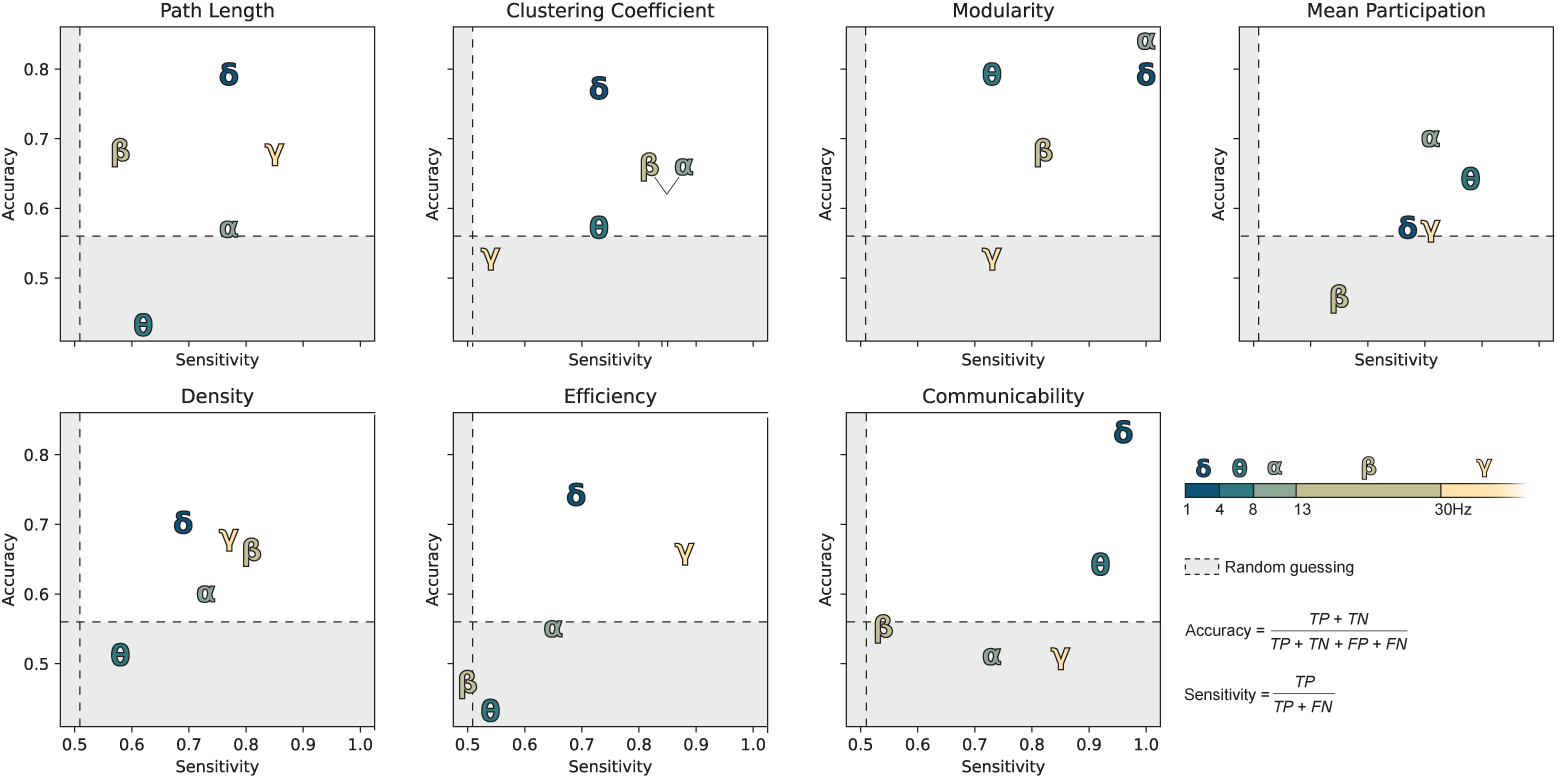
Topological-feature accuracy and sensitivity (50/50 split) across bands: delta (*δ*), theta (*θ*), alpha (*α*), beta (*β*), gamma (*γ*). Shaded area shows random-guess range based on class ratio.

We applied the same pipeline described just above for each dataset. Moreover, we extend our modelling strategy by considering separate frequency and band datasets, i.e. *TF*_1_, *TF*_2_, …, *TF*_40_ and *TF_δ_*,…, *TF_γ_*, as a parameter to be optimised. This allows us to extract the best predictive frequency and band by considering each dataset as a parameter in the optimisation process. Note that we consider both frequencies and bands separately to highlight if there is an interest in averaging features across bands or not to further summarise information.

As our classification model, we use CatBoost [69, 70], a gradient boosting method that builds decision trees sequentially, enhancing model generalisation. We chose a boosting-based method for their robustness to overfitting, which is crucial in our scenario with potential noise, as we are not thresholding the dwPLI connectivity matrices. Catboost was implemented using the *catboost* package (https://github.com/catboost). Model interpretability was achieved with Shapley values [29], a concept from game theory which attribute a quantitative contribution to every feature and thereby identify patient-specific determinants of outcome; details follow in the next section.

#### Explainability and Shapley values

Explainability clarifies how a model reaches its predictions, which is vital in clinical contexts where decisions directly affect patient care [71, 72, 73]. Complex ML systems may achieve high accuracy, yet clinicians hesitate to trust “black boxes” [14]. Transparent reasoning lets practitioners understand how a model makes its conclusions [20] For example, when predicting patient survival based on EEG functional connectivity, it is important to know which topological features or electrodes influence the model’s prediction the most. As such, a clinician can evaluate whether the model output aligns with their knowledge, helping to make better decisions [21, 22] and bridging qualitative expert evaluations [74, 4, 3, 75] with quantitative machine learning assessments [2, 9, 10, 13]. Furthermore, explainable methods provide error analysis, that is, which features contributed the most to incorrect predictions, and ensure accountability by illustrating the decision process [20, 21, 22, 15, 16, 17, 18], as illustrated in Figure 1.

We quantified feature contributions with Shapley values [29] implemented in the SHAP framework (SHapley Additive exPlanations, see [30] for more mathematical details). SHAP assigns each input a local importance score by comparing the full-model prediction with predictions obtained after withholding that feature. For functional connectivity data from EEG, this means understanding which characteristics (e.g., specific topological features or electrode interactions) are most influential for predicting patient outcomes [32]. As an illustrative example, consider a scenario in which the model predicts survival based on patterns in EEG functional connectivity. Say that the model initially predicts a high probability of non-survival based on some EEG connectivity dataset, and we systematically withhold information about specific topological features or electrode interactions; then we can observe how the prediction shifts. For instance, removing connectivity data between frontal and parietal electrodes might increase the survival probability prediction, suggesting that these region interactions are important in the original assessment of non-survival. As such, we can isolate and understand the influence of individual EEG features on each patient classification. To move beyond these insights at the instance level, one common approach is to aggregate SHAP values. However, doing so yields an “average local” importance map. This can inflate scores for correlated variables [76], which can limit feature selection or interpretation [46]. An average locality also does not reflect global model performance tied directly to its overall predictive performance as suggested in [77].

To obtain a dataset-level view we computed Shapley Additive Global importancE (SAGE) values [31]. SAGE measures the changes in the overall predictive loss when a feature (or set of features) is removed, thereby capturing its global contribution to the model’s accuracy. SAGE values also reduce the effect of correlated features on explainability by assigning importance scores relative to their correlation and impact on prediction. It is important to note that both SHAP and SAGE values face challenges-such as problems with correlated features (see [46]) - and each method has its inherent limitations. However, they capture different aspects of feature importance. Using both SHAP and SAGE provides complementary perspectives: instance-specific and global insights. This helps reduce the risk of overinterpreting a single metric, a concern called ’magical thinking’ in data science [78].

Exact Shapley computation is combinatorially costly, so we employed efficient estimators from the *shap* package (https://github.com/shap): the classic sampling method for small input sets and KernelSHAP for larger ones [30]. Global SAGE scores were obtained with the *sage* library (https://github.com/iancovert/sage) from the original paper [31].

## Results

### Univariate performances of topological features

We first assessed the baseline discriminative power of each topological feature—clustering coefficient, characteristic path length, modularity, participation coefficient, global efficiency, edge density, communicability—computed from 1–40 Hz dwPLI networks and their band averages. Figure 2 shows that the delta band consistently yields the highest accuracy across features, except for a narrow advantage of the participation coefficient and modularity in other bands. Performances of alpha, theta, beta, and gamma bands vary widely. Using the dataset selector from our pipeline, we then chose the single frequency or band maximising validation accuracy–sensitivity. For every feature except the participation coefficient, both 2 Hz and the delta band showed the highest predictive accuracy; participation peaked at 13 Hz and underperformed in the delta band. Detailed metrics are shown in the Supporting Information (S2 Table).

Recall that for dwPLI networks, edges represent phase-lag synchrony between electrode pairs. As such, a longer characteristic path length thus reflects larger average phase differences between connections. Lower global efficiency, which is associated with weaker interregional synchrony, and reduced edge density, signaling fewer active phase-lag links, both characterise poor outcomes. In contrast, high modularity identifies patients with FO and are thus characterised by segregated but internally coherent modules. Lower clustering coefficients and communicability—again prevalent in FO—indicate that there are fewer tightly interconnected nodes, suggesting a network that avoids local synchronisation and long communications.

These univariate results, highlighting the most promising frequency ranges (especially the delta band), shape the multi-feature and multi-frequency analyses that follow.

### Combining features

While the Supporting Information (S2 Table) and Figure 2 showed that 2 Hz and the delta band were strong single predictors, univariate analysis ignores interactions among metrics. Combining complementary descriptors may capture subtler network alterations relevant for survival.

Table 1 confirms that multi-feature models increase performance. Data sets *TF*_2_ and *TF_δ_* (the data sets corresponding to features at 2Hz and the delta band selected as the best data sets in all frequencies and bands, respectively; see the Section Statistical methodology for more details on the datasets) show strong predictive performance with accuracies of 0.84 and AUCs of 0.92 and 0.95. Integrating the 40 frequencies (*TF*_1:40_) preserved the 0.84 accuracy of the best univariate sets, while concatenating the band-averaged features (*TF_δ:γ_*) raised the accuracy to 0.95 and the AUC to 0.98, far exceeding the results of a single feature (shown in the Supporting Information, S2 Table). Thus, combining diverse metrics from different frequencies, from local network properties to global descriptors, provide complementary prognostic information.

**Table 1.** CatBoost performance on different datasets. Significance levels obtained by a permutation test are indicated by symbols: ^∗^ *p <* 0.05, ** *p <* 0.01, *** *p <* 0.001, ^†^ *p <* 0.10, and *^ns^* not significant (*p* ≥ 0.10). The frequency and band datasets, *TF*_2_ and *TF*_δ_, are the best predictive ones selected by the grid search as the best hyperparameters.

| Dataset | N° Features | Accuracy | AUC | PPV | NPV | Sensitivity | Specificity |
| --- | --- | --- | --- | --- | --- | --- | --- |
| $TF_2$ | 3 | 0.84** | 0.92 | 1.00 | 0.73 | 0.73 | 1.00 |
| $TF_\delta$ | 5 | 0.84** | 0.95 | 1.00 | 0.73 | 0.73 | 1.00 |
| $TF_{1:40}$ | 79 | 0.84** | 0.91 | 0.83 | 0.86 | 0.91 | 0.75 |
| $TF_{\delta:\gamma}$ | 24 | 0.95*** | 0.98 | 1.00 | 0.89 | 0.91 | 1.00 |

Figure 3 details why the combined model excels. In these plots (B and C from Figure 3, features are ranked according to their impact on model predictions, with the most influential features appearing at the top. Each point on the plot represents a patient, colour coded to indicate the feature value’s effect: red for higher values and blue for lower values that either decrease this likelihood or suggest a UO; as well as directionally coded to indicate the feature contribution to the prediction probability of the positive class (here FO): an increasing positive impact means a greater contribution towards the prediction probability of FO, a decreasing negative impact means a greater contribution towards the prediction probability of an UO. We provide both averaged SHAP values in a beeswarm plot and SAGE values to have both the local and global impact of each feature.

**Fig 3.**
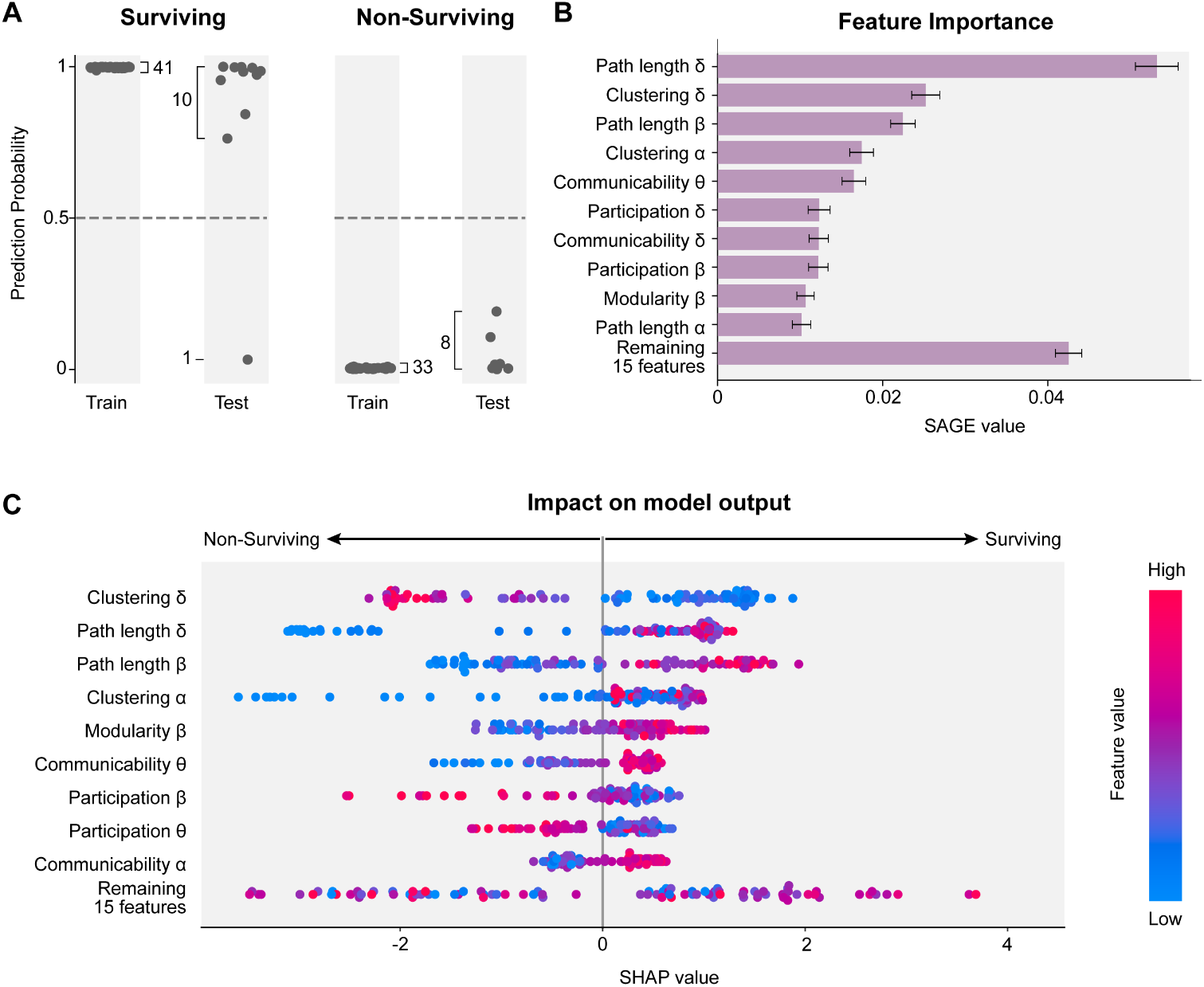
Full model explanation from the best predictive dataset, *TF_δ:γ_*, from Table 1. (A) Confidence scores were computed for each patient. A patient was predicted to be a survivor if the confidence score was above 0.5, otherwise a non-survivor. Data from patients of the train set (left subcolumns) were used to train the model. (B) Features impact on model accuracy measured by SAGE values on topological feature from *TF_δ:γ_*. (C) Features impact on prediction measured by SHAP values. Each point represents a patient. A positive impact on model output contributes to the classification of a survivor and vice versa.

For the best dataset, *TF_δ:γ_*, with 24 features selected from 35, we see from Figure 3 that SAGE identifies delta-band path length as the top global contributor, followed by delta-band clustering coefficient. SHAP assigns them the opposite order, underscoring their complementary perspectives. Low delta-band clustering and long delta-band paths favour FO. Crucially, the observed directionality of each predictor remains the same in both univariate and multivariate settings—for instance, if a low value of one feature was associated with FO in the univariate analysis, it is also the case in the multivariate model. Additional important predictors include beta-band path length and alpha-band clustering (low values linked to UO), plus delta-band participation coefficient and communicability, indicating that integration and segregation across multiple bands jointly shape outcome. The theta band also contributed through communicability which may reflect the essential role of theta rhythms in mediating communication across large-scale brain networks, which is critical for cognitive functions and may influence recovery potential [79]. In contrast, gamma-band features carry little weight, suggesting high-frequency activity is less informative in early coma, where the functionality of the basic neural network is more predictive [80] In essence, while the univariate results highlighted the role of the delta band as a strong independent predictor, the multifeature approach shows a subtle interplay among features from different frequency ranges, providing a more robust and comprehensive characterisation of functional connectivity.

Overall, combining weighted-network metrics across bands not only improves accuracy but also supports the notion that distinct frequency bands capture complementary aspects of brain activity that single-band or binarised methods might overlook. Delta-band topology—especially prolonged paths and reduced clustering—remains pivotal, yet adding alpha, beta, and theta bands descriptors refines prognoses without sacrificing interpretability, paving the way for patient-specific explanations in the next section.

### Explainable diagnosis of missclassified patients

Despite the high overall accuracy of our models, some patients were misclassified. To understand these misclassifications, we use our explainable framework with Shapley values to investigate the individual contributions of features to each patient’s prediction. This explainable framework allows us to understand not only which features influenced the model’s decisions but also how they contributed to incorrect classifications.

We focus on two illustrative cases where topological features from all bands are used (i.e., with *TF_δ:γ_*): a surviving patient (FO) who was incorrectly classified as non-surviving (UO) and a non-surviving patient (UO) who was incorrectly classified as surviving (FO). The Shapley bar plots for these patients are presented in Figure 4.

**Fig 4.**
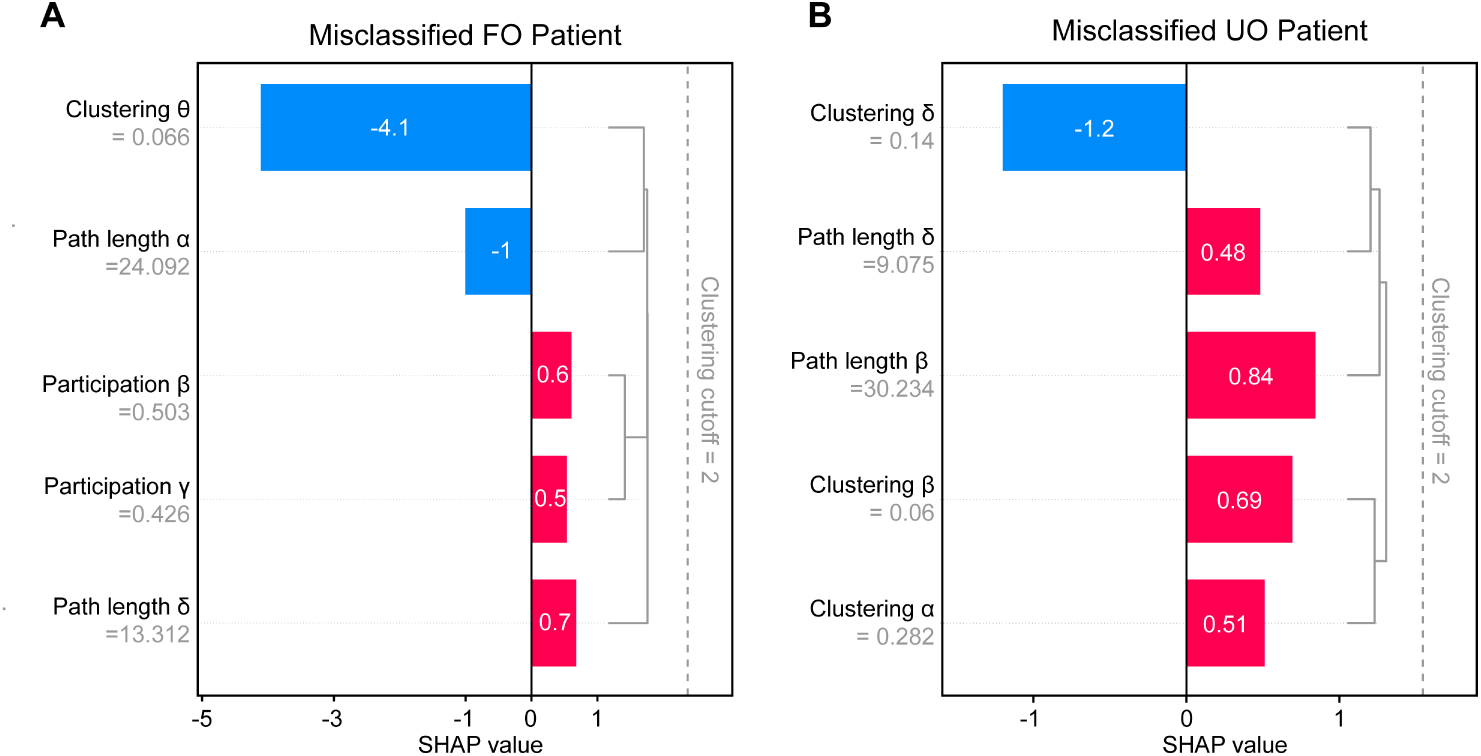
Features impact on prediction measured by SHAP values on topological features from *TF_δ:γ_* on misclassified patients. (A) Local SHAP plot representing the impact of each features on a FO patient being wrongly classified as UO. (B) Local SHAP plot representing the impact of each features on a UO patient being wrongly classified as FO.

Each bar in the plots represents a topological feature with the length indicating the magnitude of its contribution to the prediction. Positive values push the prediction towards FO (positive impact on the probabilistic decision, corresponding to the red bars), while negative values push it towards UO (negative impact on the probabilistic decision, corresponding to the blue bars). For the FO patient predicted UO, negative SHAP attributions dominated: notably, a low theta-band clustering coefficient—associated with UO—pushed the score below the survival threshold. The patient’s connectivity profile thus resembled the non-survivor group. Conversely, the UO patient erroneously classified FO showed strong positive attributions, driven by high path lengths in delta and beta bands, features linked to favourable outcomes.

By systematically examining these outlier cases, we demonstrate how Shapley-value explanations can guide further clinical investigation. Importantly, none of these interpretability methods altered the model’s raw accuracy; instead, they provided transparency to clinicians. Patient-specific explanations highlight ambiguous EEG characteristics that may benefit from additional biomarkers or clinical assessments to refine prognostic certainty and consequently further improve predictions.

## Discussion

In this work, weighted EEG functional-connectivity networks were analysed across 1–40 Hz and summarised with seven topological measures. Combining frequencies at the band level produced the highest accuracy (0.95) while retaining explainability through SHAP and SAGE analyses that highlighted long path length and low clustering in the delta range as decisive markers. Patient-specific explanations clarified why borderline cases were misclassified and showed how cross-band interactions refine predictions beyond any single metric.

Functional connectivity features and long range connections at different frequency bands has been investigated in different varieties of unconscious states for identifying markers of consciousness levels and improve prognostication [2, 9, 10, 81]. While power spectra analysis suggests a distinctive role of delta, theta and alpha frequency range for discriminating levels of consciousness in minimally consciousness level patients, vegetative patients and healthy participants [2], functional connectivity based on phase synchronisation in the delta range provided accurate stratification of consciousness levels in vegetative and minimally consciousness state patients [9]. Across varieties of physiological and clinical consciousness alterations, including anaesthesia, sleep, and disorders of consciousness patients, functional connectivity based on weighted symbolic mutual information emerged as the most robust index for stratifying consciousness levels [81]. Our results, highlighting a distinct role of the delta band in the prognosis of the outcome of comatose patients, complement these previous findings by suggesting that functional connectivity organization in the delta range may constitute part of the precondition for consciousness recovery in comatose patients. An alternative and not exclusive argument for interpreting our results in relation to previous literature is that in patients with favourable outcomes recorded during the first day of coma, some residual level of consciousness may remain undetected by standard clinical examinations (based on the Glasgow coma scale [47] and the FOUR score [48]). If this alternative hypothesis was valid, our results would align and expand previous observations on functional connectivity in the delta range being a robust marker for discriminating consciousness levels even during the very first day of coma.

Taken together, the present findings expand the prior literature by demonstrating that the early-coma delta-band topology, captured with weighted networks and interpreted through explainable AI, offers a transparent and statistically robust prognostic tool. The framework’s ability to identify feature interactions indicates potential directions for multimodal integration, linking EEG with imaging or biomarkers, to further personalise outcome prediction and guide targeted interventions.

### Explainable classification by combining network features from different band

[9] and [13] showed that topological features extracted from binarised EEG networks highlight brain organisation, but thresholding can discard signal. Adapting the workflow of [13] to weighted graphs, we achieved higher accuracy.

In univariate tests, the most discriminative frequencies are predominantly in the delta band, specifically at 2Hz. Favourable outcome (FO) corresponded to longer path length and higher modularity but lower clustering and participation coefficients (respectively at 2 Hz, 2 Hz, 2 Hz, 13 Hz). These findings echo the 10 Hz results of [13], indicating that FO brains retain densely connected meso-communities with weak local cohesion. The participation coefficient itself was uninformative in the delta band, suggesting similar inter-module hub strength in both groups. Other metrics—edge density, global efficiency, communicability—were again most predictive at 2 Hz: lower values signalled FO, implying globally sparse, inefficient, and weakly redundant networks. Although seemingly counter-intuitive, this inefficiency refers only to graph-theoretic measures, not clinical function.

Multi-feature models substantially improved performance. Integrating metrics within single delta frequencies (*TF_δ_*) or all 40 frequencies (*TF*_1:40_) sustained 0.84 accuracy, yet averaging by band and concatenating (*TF_δ:γ_*) raised accuracy to 0.95 with an AUC of 0.98 while using just 24 parameters. Averaging over bands thus reduces noise, improves interpretation, and enhances statistical significance (see table 1 and the Supporting Information, S2 Table). Our *TF_δ:γ_* model (accuracy = 0.95, AUC = 0.98, sensitivity = 0.91, specificity = 1.00) clearly surpasses earlier EEG-based prognostic tools. [12] combined functional-connectivity and non-coupling metrics to reach sensitivity = 0.73, specificity = 1.00, AUC = 0.92. [13] reported maximal AUCs of 0.77 (clustering), 0.72 (path length) and an overall best accuracy of 0.71 when using modularity time-variance (PPV = 0.67, specificity = 0.32). [9] obtained AUC = 0.83 and 79 % accuracy with alpha-band participation coefficients, while [11] showed a modest improvement over clinical scores (|t| = 2.98, p = 0.007). These head-to-head figures underscore the benefit of combining weighted-network features across bands while preserving model transparency through concise explanation summaries (see Figure 3).

Furthermore, the explainable nature of our classification method allows us to reveal the decision process for each patient and show which features were important in making the decision, a level of interpretability that is rarely done in comparable studies such as [13], [11] and [12]. Understanding why a patient was misclassified can offer insight into atypical cases and, consequently, change treatment plans. This aligns with the clinician-centred view that explanations must support rationalisation steps after a prediction, allowing an expert to validate the output against domain knowledge [14]. This is particularly important because, as illustrated in Figure 4, some patients exhibit features that align more closely with the opposite group. These borderline cases might benefit from additional expert evaluation, such as taking into account other clinical variables, highlighting the need to integrate clinical judgment with model predictions.

In addition, this analysis highlights the importance of considering individual patient profiles rather than relying only on aggregate data. Misclassified patients might possess unique physiological or pathological characteristics not captured entirely by EEG data, even less by topological features. Explainable insights into these individual profiles can guide the collection of additional data modalities. This is especially the case for the non-surviving cases that had features similar to the surviving group, which could provide valuable information on the underlying mechanisms of recovery or deterioration in comatose patients. Such patient-specific explanations may also reveal latent subgroups, informing both prognosis and tailored interventions. As such, additional clinical variables, such as patient history or biomarkers, can potentially improve the overall accuracy of the model, reduce misclassifications [17], but also contribute to personalised medicine approaches in critical care settings [19, 23]. By integrating additional clinical data, our explainability framework can extend to causal or symbolic explanations, further bridging the gap between black-box predictions and actionable clinical insights [82].

### Limitations of our method

The limitations in the data itself are inherited from [13]. That is, its dependence on an experienced electrophysiologist to visually inspect and filter out noisy data in a supervising data rejection scheme. Moreover, we must note that muscle artefacts lead to the exclusion of 28% of patients due to the increased sensitivity of connectivity metrics to noise.

In studies such as [9] or [13], topological features are derived from the thresholded and binarised weighted networks to remove potential noise from functional connectivity. Here instead, we derive topological features from the weighted networks directly. The downside is this approach keeps more of the noise, and thus can be more sensitive to noisy weights. However, the dwPLI is known to be robust to noise [37], and therefore when combined with cross validation this risk is mitigated. In fact, we see that we obtain better predictions compared to [13].

A limitation of explainability is related to its benefit in clinical uses. In this context, Shapley-based explanations are used for normative evaluations [83, 84], such as helping a human to evaluate a specific decision made by a model. Some studies showed that an explainable framework did not present significant evidence that its users were better able to assess the accuracy of the predictions [85]. For example, [86] showed that the increased transparency of an explained model hindered people’s ability to detect when the model made a substantial mistake and correct for it. However, we must note that, in the clinical context, the impact of explainability on trust and transparency is mixed, as extensively described in [87]. [88] and [15] revealed that physicians who could explain the algorithm’s decision to a patient were more inclined to follow the recommendation. Trustworthiness is also mixed as some studies show improvement [18, 16, 17], while others show no significant improvement from adding explanations to the model predictions, as seen in [89]. However, they clarify that it might be due to explanations being almost always similar to clinician expectations. [18] concluded that, although inconsistency between the model’s predictions and the clinicians’ opinion could severely affect clinicians trust, explanations overall improved perceived trustworthiness.

## Conclusion

In this study, we introduced an explainable machine learning pipeline for the early prediction of the outcomes of comatose patients after cardiac arrest, using weighted functional connectivity and topological network features derived from EEG data. Unlike conventional threshold-based methods, our approach preserves and exploits the continuous nature of brain synchrony, capturing subtleties that binarised networks might overlook. By focussing on multiple frequencies, from 1 to 40 Hz, and employing Shapley values (both local SHAP and global SAGE) for interpretability, we demonstrated that the combination of frequency-specific features substantially improves classification performance.

Our results reveal that low-frequency bands (especially the delta band at 2 Hz) carry strong prognostic information, extending earlier work [9, 13] that focused on alpha and theta bands by highlighting the added value of lower frequencies. In addition, the explainable framework shines a light on individual misclassifications, showing that patient-level differences can be scrutinised to understand why an EEG profile appears more characteristic of a particular outcome group. This transparency is critical to building clinician trust and potentially guiding more personalised patient care.

By integrating weighted network topology, cross-frequency analysis, and interpretable AI methods, this pipeline offers a powerful alternative to existing black-box solutions. Future studies can build on this work by automating artifact removal, exploring more advanced weighted network denoising, and incorporating clinical and biomarker data for holistic patient evaluations. Ultimately, our findings show that the use of explainable, multifrequency EEG connectivity models can provide highly accurate and clinically interpretable early outcome predictions, an essential step toward improving critical care decision-making for comatose patients.

## Supporting information

**S1 Text. Weighted topological features mathematical description.**

**S1 Table. Weighted topological feature list and references.**

**S2 Table. Performance of single topological features.**

## Data availability

The data included in this study are not publicly available due to the sensitive nature of the clinical EEG recordings and restrictions imposed by Swiss ethical and legal frameworks. According to the ethics approval (PB 2016-00530, CER-VD, 23/05), these data cannot be shared outside the approved research consortium. For more information about the ethical framework governing these data, please contact the *Commission cantonale d’éthique de la recherche sur l’être humain du Canton de Vaud* (CER-VD), which oversees research ethics and compliance for this cohort but does not distribute or authorize data access. The analysis code used for data processing and model training is available from the authors upon reasonable request.

## Acknowledgments

We thank Thomas Kusterman for data curation and preprocessing. We thank all the clinicians involved in the study: Matthias Haenggi, Rebekka Kurman, Frederic Zubler, Mauro Oddo, Andrea Rossetti. We thank all EEG technicians from the involved hospitals for invaluable help with EEG recordings. We thank Christine Staehli, R.N., and Daria Solari, M.D., from the Lausanne University Hospital, Dragana Viceic, M.D. PhD, and Marco Rusca, M.D., from the Sion Hospital, Ettore Accolla, M.D., and Sebastien Doll, M.D., from the Fribourg Hospital for providing clinical information for some of the patients. We are indebted to Rupert Ortner and Christoph Guger for technical support.

This work was supported by the European Research Council under Grant CoG 2015-682172NETS to S.C.O, within the Seventh European Union Framework Program, the Swiss NSF (grant 32003B 212981) to M.D.L and the Eurostars project E!3489 to M.D.L.

