## Supplementary materials for "Explainable machine learning on weighted connectivity networks across frequencies for outcome prediction in comatose patients"

### S1 Text: Weighted topological features.

This descriptive section will mainly follow the work and descriptions of [1], [2], [3], [4], [5], [6], [7] as well as the summary from [8].

#### Basic features: direct-link measures

In the weighted case, each existing link $(i, j)$ is associated with a weight $w_{ij}$, normalized so that $0 \leq w_{ij} \leq 1$. The *weighted edge density* $D^{\mathrm{w}}$ is defined as

$$D^{\mathrm{w}} = \frac{\sum_{i,j\in[n]} w_{ij} A_{ij}}{n(n-1)\max_{i,j\in[n]}(w_{ij})}.$$

Here, the denominator $n(n-1)\max_{i,j\in[n]}(w_{ij})$ corresponds to the highest possible sum of edge weights in a fully connected network of $n$ nodes, where each of the $n(n-1)/2$ edges is assigned the maximum observed weight. By summing over all ordered pairs $(i, j)$, we naturally count each undirected link twice, maintaining consistency with the way $A_{ij}$ is defined. This normalisation ensures that $D^{\mathrm{w}}$ ranges from 0 to 1, effectively capturing the proportion of the maximum attainable weighted connectivity that the network achieves. Note that if all weights have a value of 1, then the subsequent graph is a binary graph.

Recall that for a binary (unweighted) network, the degree of a node $i \in [n]$ is

$$k_i = \sum_{j\in[n]} A_{ij}.$$

In a weighted network, the analogous notion is the *weighted degree*, also referred to as the node strength. For node $i \in [n]$, the weighted degree $k_i^{\mathrm{w}}$ is defined as

$$k_i^{\mathrm{w}} = \sum_{j\in[n]} w_{ij}.$$

This weighted degree $k_i^{\mathrm{w}}$ quantifies not only how many links connect to node $i$, but also the intensity of those connections.

#### Integration features: distance-based measure

##### Node distance

Now, we define the notion of shortest path lengths (node distances) in a network. In a binary network, it is defined as

$$d_{ij} = \sum_{(u,v)\in g_{i\leftrightarrow j}} A_{uv},$$

where $g_{i\leftrightarrow j}$ is the shortest path (geodesic) between $i$ and $j$, for $i, j \in [n]$. In a weighted network, however, the notion of distance depends on the nature of the network. Following [5], there are two distinct definitions depending on its nature: one for transportation (remoteness) networks and one for communication (proximity) networks. Consider an undirected weighted network with normalised weights $w_{ij}$ ($0 \le w_{ij} \le 1$). The shortest path between two nodes $i, j \in [n]$, denoted $g_{i\leftrightarrow j}$, is the path that minimises the total traversal cost defined according to the type of network. For transportation networks, the weighted shortest path length $d_{ij}^{\mathrm{tr},w}$ is defined as the minimal sum of weights along any path from node $i$ to node $j$, for $i, j \in [n]$:

$$d_{ij}^{\mathrm{tr},w} = \sum_{(u,v)\in g_{i\leftrightarrow j}} w_{uv}.$$

Here, the weight $w_{uv}$ can be viewed as a physical distance or cost. Thus, a stronger connection is represented by a lower weight, making shorter paths more efficient in a transportation context. For communication networks, it is the inverse: the strength of a connection is proportional to its weight. Thus, the shortest path length is defined as the minimal sum of the inverse weights along any path:

$$d_{ij}^{\mathrm{com},w} = \sum_{(u,v)\in g_{i\leftrightarrow j}} \frac{1}{w_{uv}}.$$

In both definitions, if no path exists between $i$ and $j$, we set $d_{ij} = \infty$.

##### Characteristic path length

For a binary (unweighted) network, the characteristic path length $L$ is the average shortest path length over all pairs of distinct nodes:

$$L = \frac{1}{n} \sum_{i\in[n]} \frac{\sum_{j\in[n], j\neq i} d_{ij}}{n-1},$$

where $d_{ij}$ is the binary shortest path length between $i$ and $j$. For weighted networks, the weighted characteristic path length $L^{\mathrm{w}}$ is defined similarly, using $d_{ij}^{\mathrm{w}}$, the shortest weighted path length (defined as either $d_{ij}^{\mathrm{tr},w}$ or $d_{ij}^{\mathrm{com},w}$, depending on the network context):

$$L^{\mathrm{w}} = \frac{1}{n} \sum_{i\in[n]} \frac{\sum_{j\in[n], j\neq i} d_{ij}^{\mathrm{w}}}{n-1}.$$

##### Global efficiency

Network efficiency [9, 4] measures how efficiently information is exchanged in a network. For a binary network, the global efficiency $E$ is the average of the inverse shortest path lengths:

$$E = \frac{1}{n} \sum_{i\in[n]} \frac{\sum_{j\in[n], j\neq i} d_{ij}^{-1}}{n-1}.$$

For weighted networks, using the shortest weighted paths (e.g., $d_{ij}^{\mathrm{com}}$ in a communication network), the weighted global efficiency $E^{\mathrm{w}}$ is:

$$E^{\mathrm{w}} = \frac{1}{n} \sum_{i\in[n]} \frac{\sum_{j\in[n], j\neq i} (d_{ij}^{\mathrm{w}})^{-1}}{n-1}.$$

#### Average communicability

The notion of *communicability* has first been introduced for binary networks in [10] as a more general measure of connectedness. In fact, communicability was generally considered the shortest path that connects two nodes. However, in real-world networks, communication between two nodes can happen along non-shortest paths. Moreover, taking the shortest paths as a measure of communicability does not reflect the impact of *structural bottlenecks*, such as bridges, on the overall communicability of a network. As such, it is sensible to propose a measure, here *communicability*, that also accounts for other paths one can take to travel from one node to another.

*Communicability* between node $i$ and $j$, for $i \neq j$, written $G_{ij}$, in the sense of [10], makes longer walk have lower contribution to the measure and, as such is defined as follows in [10] for $i \neq j$

$$G_{ij} = \frac{1}{s!} P_{ij}^{(s)} + \sum_{k>s} \frac{1}{k!} W_{ij}^{(k)}, \tag{1}$$

where $P_{ij}^{(s)}$ is the number of shortest paths between the nodes $i$ and $j$ having length $s$, and $W_{ij}^{(k)}$ is the number of walks connecting $i$ and $j$ of length $k > s$. Here, longer walks of length $k$ are penalised by a factor of $1/(k!)$. Using standard results between the powers of the adjacency matrix and the number of walks in the network, we can express (1) as follows, for $i \neq j$,

$$G_{ij} = \sum_{k=0}^{\infty} \frac{\left(A^k\right)_{ij}}{k!} = \left(e^A\right)_{ij}.$$

We can simplify this representation further by using the spectral decomposition of the adjacency matrix $A := \{A\}_{1 \leq i,j \leq n}$. Indeed, we have $A = \Phi \Lambda \Phi^\top$, where $\Phi$ is the orthogonal matrix of eigenvectors $\varphi_j$ and $\Lambda = \operatorname{diag}(\lambda_1, \ldots, \lambda_n)$ contains the eigenvalues $\lambda_j$. Applying the matrix exponential, we obtain $e^A = \Phi e^\Lambda \Phi^\top$. Consequently, the $(i, j)$-th entry of $e^A$, which represents the communicability $G_{ij}$, for $i \neq j$, is given by

$$G_{ij} = \left(e^A\right)_{ij} = \sum_{p=1}^{n} \varphi_p(i) \varphi_p(j) e^{\lambda_p}$$

thereby expressing communicability in terms of the graph's spectral properties through the eigenvalues $\lambda_p$ and the corresponding orthonormal eigenvectors $\varphi_p$. It can be shown that this expression can be further reduced, using the graph spectrum [11, 12], to

$$G_{ij} = \sum_{p=1}^{n} \varphi_p(i) \varphi_p(j) e^{\lambda_p},$$

where $\varphi_p(i)$ is the $i$th element of the $p$th orthonormal eigenvector of the adjacency matrix associated with the eigenvalue $\lambda_p$. More compactly, it can be defined as

$$\exp(A)_{ij},$$

for $i \neq j$, as mentioned in [13]. They further adapted the notion to weighted proximity networks. However, adapting it directly using $\exp(A)$ creates a similar problem observed in spectral clustering of nodes, namely that the size of a cluster was more influenced by the number of nodes than the total weight of connections within the clusters [14]. As in spectral clustering with normalised Laplacian, communicability adapts to weighted networks using degree correction and is thus defined as

$$G_{ij} = \left(\exp\left(D^{-1/2} A D^{-1/2}\right)\right)_{ij},$$

for $i \neq j$, where the diagonal degree matrix $D \in \mathbb{R}^{n \times n}$ has the form $D := \operatorname{diag}(k_i^w)$. As a topological feature, we thus use the average communicability in the network.

#### Segregation features: clustering-based measures

##### Clustering Coefficient

A standard approach to investigating the presence of communities or cohesive groups in a network is to measure its clustering coefficient [15, 3, 16, 17]. The clustering coefficient quantifies the tendency of nodes to form tightly-knit groups of interconnected nodes, commonly measured by the prevalence of triangles in the network. For a binary (unweighted) network with $n$ nodes, we define the number of triangles around a node $i$, for $i \in [n]$, as

$$t_i = \frac{1}{2} \sum_{j,h \in [n]; j,h \neq i} A_{ij} A_{ih} A_{jh}.$$

A triangle is thus formed whenever three nodes all share mutual connections. The local clustering coefficient of node $i$ in a binary network is given by [15]:

$$C_i = \frac{2t_i}{k_i(k_i - 1)},$$

By averaging over all nodes, we obtain the global clustering coefficient as

$$C = \frac{1}{n} \sum_{i \in [n]} C_i.$$

In weighted networks, the concept is adapted to incorporate edge weights. We define the weighted geometric mean of the triangles around node $i$ [18] as

$$t_i^{\mathrm{w}} = \frac{1}{2} \sum_{j,h \in [n]; j,h \neq i} (w_{ij} w_{ih} w_{jh})^{1/3}.$$

Here, the weights $w_{ij}$ are normalized by the maximum weight in the network. The weighted local clustering coefficient of node $i$ is then

$$C_i^{\mathrm{w}} = \frac{2t_i^{\mathrm{w}}}{k_i(k_i - 1)},$$

and the global weighted clustering coefficient is

$$C^{\mathrm{w}} = \frac{1}{n} \sum_{i \in [n]} C_i^{\mathrm{w}}.$$

The global weighted clustering coefficient defined here, from [18] is adapted to measure local cohesiveness in weighted networks as, in essence, it depends on the ratio of triangles and their total relative link weight with respect to the strength of the node [7]. Other definitions are reviewed in [17]. Note that the clustering coefficient has also been adapted to account for negative weights [19] and directed networks [20]. However, such methods are not investigated here, as we consider weighted matrices which are positive and undirected.

##### Modularity

The detection of community structure in networks can be addressed with *modularity*, a measure that quantifies how well a network can be partitioned into densely connected groups (modules) with sparser connections between them [21, 2], known as an assortative behaviour. More particularly, the modularity $Q$ of a network measures the segregation of different communities. For binary networks fully subdivided into a set of non-overlapping modules $Z$, the modularity is given by [1, 2]

$$Q = \frac{1}{l} \sum_{i,j \in [n]} \left( A_{ij} - \frac{k_i k_j}{l} \right) \delta_{z_i z_j},$$

where $l = \sum_{i,j\in[n]} A_{ij}$ is the total count of links (counting each undirected link twice), $k_i$ is the degree of node $i$, $z_i \in [Z]$ is the module of node $i$, and $\delta_{z_i z_j} = 1$ if $z_i = z_j$ and 0 otherwise. As such, the modularity $Q$ represents the fraction of links within a given community minus its expected fraction of links that are drawn at random. Positive $Q$ indicates a good community division, and is more defined as $Q$ converges to 1. For weighted networks, the modularity can be adapted by replacing the binary degrees with weighted degrees $k_i^{\mathrm{w}} = \sum_{j\in[n]} w_{ij}$ and the total link count with the total weight $l^{\mathrm{w}} = \sum_{i,j\in[n]} w_{ij}$

$$Q^{\mathrm{w}} = \frac{1}{l^{\mathrm{w}}} \sum_{i,j\in[n]} \left( w_{ij} - \frac{k_i^{\mathrm{w}} k_j^{\mathrm{w}}}{l^{\mathrm{w}}} \right) \delta_{z_i z_j}.$$

A higher $Q^{\mathrm{w}}$ indicates a stronger community structure when considering both the presence and intensity of connections. Efficient methods such as the one by [22] can be used to estimate weighted modularity.

**Participation Coefficient**

The participation coefficient [23] measures how well connected a node is to different modules of the network. It evaluates whether a node's links (or strengths in the weighted case) are evenly distributed among all modules or concentrated within a single module. For a binary network, the participation coefficient of a node $i \in [n]$ is:

$$P_i = 1 - \sum_{z\in Z} \left( \frac{k_i(z)}{k_i} \right)^2,$$

where $k_i(z)$ is the number of links between node $i$ and module $z$, and $k_i$ is the total degree of node $i$. For weighted networks, this definition is naturally extended by using weighted degrees:

$$P_i^{\mathrm{w}} = 1 - \sum_{z\in Z} \left( \frac{k_i^{\mathrm{w}}(z)}{k_i^{\mathrm{w}}} \right)^2,$$

for $i \in [n]$, where $k_i^{\mathrm{w}}(m)$ is the sum of the edges weights between node $i$ and all nodes in module $m$, and $k_i^{\mathrm{w}}$ is the weighted degree of node $i$. The participation coefficient, in its binary or weighted form, helps identify nodes that act as connectors between multiple communities versus those that are predominantly embedded within a single module.

**Table S1.** Summary of topological features adapted for weighted networks

| Feature Name | Interpretation | Mathematical Intuition | References |
|---|---|---|---|
| Clustering Coefficient | Measures the tendency of nodes to cluster together, indicating local connectivity | Average strength of connectivity between a node's neighbours, considering edge weights | [5] |
| Path Length | Represents the average shortest path length between all pairs of nodes | Calculated using inverse edge weights to represent distances; averages minimal weighted path lengths | [5] |
| Modularity | Measures the division of the network into communities, indicating segregation | Quantifies the density of intra-module links compared to inter-module links, adapted for weighted networks | [22, 24] |
| Participation Coefficient | Assesses how nodes connect across different modules, indicating integration capacity | Based on weighted degrees; measures the distribution of a node's links among modules | [23] |
| Global Efficiency | Quantifies the network's ability to transfer information efficiently | Average inverse shortest path length between all node pairs, using weighted distances | [4] |
| Edge Density | Represents the general connectivity level of the network | Ratio of total edge weights to the maximum possible total weight | [25] for an EEG example |
| Communicability | Considers all possible paths between nodes, indicating robustness of information transfer | Sum over all paths, weighted by path lengths; higher for nodes connected through many short paths | [13] |

**Table S2.** CatBoost performance on different features. Significance levels obtained by a permutation test are indicated by symbols: $^{*}$ $p < 0.05$, $^{**}$ $p < 0.01$, $^{***}$ $p < 0.001$, $^{\dagger}$ $p < 0.10$, and $^{ns}$ not significant ($p \geq 0.10$). The frequency and band datasets, $TF_2$ and $TF_\delta$, are the best predictive ones selected by the grid search as the best hyperparameters.

| Feature | Hz/Band | Accuracy | AUC | PPV | NPV | Sensitivity | Specificity |
|---|---|---|---|---|---|---|---|
| Density | 2Hz | $0.79^{*}$ | 0.89 | 0.82 | 0.75 | 0.82 | 0.75 |
| Density | $\delta$ | $0.79^{*}$ | 0.86 | 0.82 | 0.75 | 0.82 | 0.75 |
| Path Length | 1Hz | $0.58^{ns}$ | 0.72 | 0.64 | 0.50 | 0.64 | 0.50 |
| Path Length | $\delta$ | $0.68^{ns}$ | 0.79 | 0.73 | 0.62 | 0.73 | 0.62 |
| Efficiency | 2Hz | $0.79^{*}$ | 0.80 | 0.77 | 0.83 | 0.91 | 0.62 |
| Efficiency | $\delta$ | $0.74^{*}$ | 0.81 | 0.80 | 0.67 | 0.73 | 0.75 |
| Clustering | 2Hz | $0.84^{**}$ | 0.78 | 0.83 | 0.86 | 0.91 | 0.75 |
| Clustering | $\delta$ | $0.84^{**}$ | 0.85 | 0.90 | 0.78 | 0.82 | 0.88 |
| Modularity | 2Hz | $0.84^{**}$ | 0.98 | 1.00 | 0.73 | 0.73 | 1.00 |
| Modularity | $\delta$ | $0.79^{**}$ | 0.73 | 0.73 | 1.00 | 1.00 | 0.50 |
| Participation | 13Hz | $0.58^{ns}$ | 0.50 | 0.60 | 0.50 | 0.82 | 0.25 |
| Participation | $\alpha$ | $0.53^{ns}$ | 0.66 | 0.57 | 0.40 | 0.73 | 0.25 |
| Communicability | 2Hz | $0.63^{ns}$ | 0.64 | 0.70 | 0.56 | 0.64 | 0.62 |
| Communicability | $\delta$ | $0.79^{*}$ | 0.73 | 0.82 | 0.75 | 0.82 | 0.75 |
